# Characterisation of peri-implantation endometrial Treg and identification of an altered phenotype in recurrent pregnancy loss

**DOI:** 10.1101/2021.04.22.21255931

**Authors:** Ingrid Granne, Mengni Shen, Helena Rodriguez-Caro, Gurmeher Chadha, Elizabeth O’Donnell, Jan J. Brosens, Siobhan Quenby, Tim Child, Jennifer H. Southcombe

## Abstract

Recurrent Pregnancy Loss (RPL) affects 2-4% of couples, and with increasing numbers of pregnancy losses the risk of miscarrying a euploid pregnancy is increased, suggesting RPL is a pathology distinct from sporadic miscarriage that is due largely to lethal embryonic aneuploidy. There are a number of conditions associated with RPL including unspecified ‘immune’ pathologies, one of the strongest candidates for dysregulation remains T regulatory cells as depletion in the very early stages of pregnancy in mice leads to pregnancy loss.

Human endometrial Treg and conventional CD4T cells were isolated during the peri-implantation period of the menstrual cycle in normal women. We identified an endometrial Treg transcriptomic signature and validated an enhanced regulatory phenotype compared to peripheral blood Treg. Parous women had an altered endometrial Treg transcriptome compared to nulliparity, indicating acquired immunity memory of pregnancy within the Treg population, by comparison endometrial conventional CD4T cells were not altered. We compared primary and secondary RPL to nulliparous or parous controls respectively. Both RPL subgroups displayed differentially expressed Treg gene transcriptomes compared to controls. We found increased cell surface S1PR1 and decreased TIGIT protein expression by Treg in primary RPL, confirming the presence of altered Treg in the peri-implantation RPL endometrium.

## Introduction

The female reproductive tract (FRT) is a mucosal barrier tissue and like other mucosa, immune cells infiltrate the stromal layers adjacent to the epithelial surface. Throughout the hormonally controlled menstrual cycle host immunity provides protection from pathogens that may enter the FRT, however when an embryo implants into the endometrium, the immune system is temporarily modified to permit attachment and invasion of trophoblast cells from the developing conceptus. At the time of embryonic implantation NK, T, B and macrophage cells are present in the human endometrium (1). Sporadic pregnancy loss is most commonly caused by embryonic aneuploidy, but with increasing numbers of pregnancy losses the risk of losing a euploid pregnancy is increased. RPL is the consecutive loss of two or more pregnancies, before 24 weeks’ gestation (2, 3), and can affect nulliparous (primary RPL) or parous women (secondary RPL). RPL is partly associated with genetic variation (4), uterine anomalies, endocrine dysfunction, parental balanced chromosomal translocation and specific maternal autoantibodies (5), however these associated clinical factors are identified in fewer than 50% of cases. The maternal immune system is implicated in some cases of recurrent pregnancy loss (6).

NK cell densities are associated with RPL (5), and CD8T cells are also known to be phenotypically altered (7). The CD4T cell compartment (consisting of Th1, Th2, Th17 and Treg defined by cytokine production) has not been extensively studied. Of specific interest are the Treg cells, which in mice have been show to prime tolerance, as depletion during early stages of pregnancy during embryonic implantation leads to pregnancy loss (8). Murine Treg cells are also pivotal to tolerance of paternal alloantigen (9). The transcription factor *FOXP3* confers regulatory lineage commitment to Tregs, multiple mechanisms of action are known such as high CD25 expression which consumes IL-2 preventing effector T cell functions, CTLA4 driven inhibition of antigen presenting cells, and inhibitory cytokine production (IL-10, TGF-B and IL-35) (10). Decreased numbers of FOXP3+ cells are found in endometrium of patients with RPL compared to fertile controls (11), however in humans *FOXP3* can also be expressed by activated T cells and other cells (12) and the phenotype and functional nature of these cells has not been clearly elucidated.

In this study human endometrium was collected during the peri-implantation period of the menstrual cycle, from normal fertile women and from women with RPL. In addition peripheral blood samples were taken from women with RPL. Transcriptome analysis of purified Treg and Tconv (CD4T cells depleted of Treg) was performed, we compared the signature of Treg versus Tconv and compared cells between nulliparous women and those who had had a previous live birth. Phenotypic changes were found between Treg and Tconv in the peripheral blood and endometrium in RPL. Treg and Tconv were also compared between normal women and RPL.

## Results

### The endometrial CD4-T cell compartment in normal physiology

Endometrial biopsies were taken during the peri-implantation window of the menstrual cycle. Single cell suspensions were obtained and flow cytometry performed to identify the total CD4T cell population (CD45/CD3/CD4), and the Treg subpopulation (CD25hi/CD127lo/FoxP3+), a representative example is shown in figure 1a. We first analysed this compartment in fertile women (nulliparous and parous combined). CD4T cells comprise 43.0±9.5% of the total CD3 positive T cell population, which is a lower proportion that the equivalent compartment in the peripheral blood, figure 1b. Treg were 5.5±3.5% of the total CD4T cell compartment in the human endometrium, this is similar to the proportion of Treg in the CD4T compartment in the peripheral blood, figure 1c. A higher proportion of CD4T cells in the endometrium are CD45RO+ memory cells (Tconv 92.6% versus Treg 97.9%) compared to those in the circulation (figure 1d), indicating a tissue specific enrichment of memory cells warranting further analysis.

**Figure 1.**
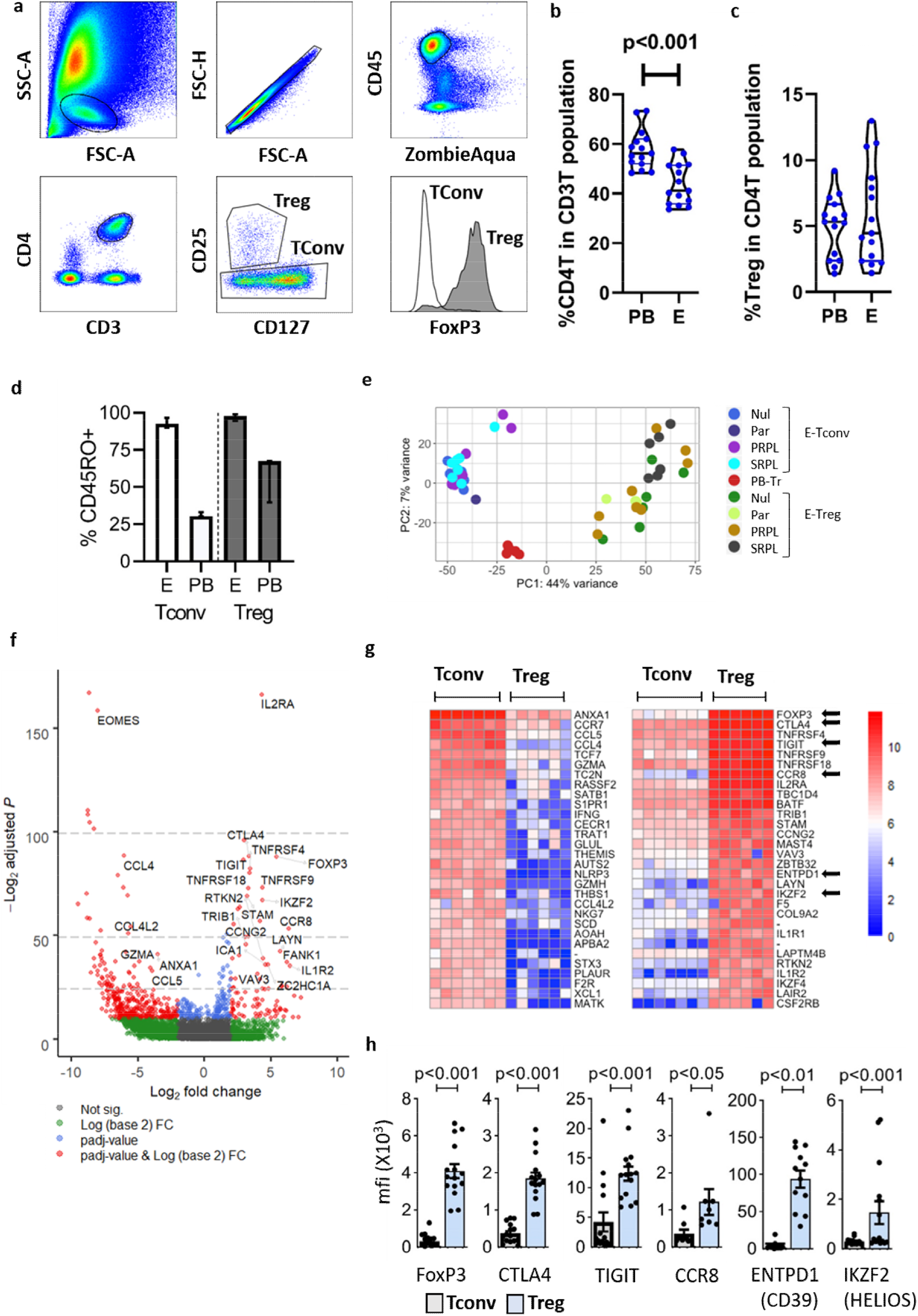
CD4T cells in the human endometrium. (a) Representative flow cytometric staining of Treg and Tconv cells in human endometrium: FSC-A vs. SSC-A and FSC-H determine lymphocyte singlet cells, ZA^lo^/CD45^+^/CD3^+^/CD4^+^ (total CD4T cells) are subpopulated CD25^hi^/CD127^lo^/FOXP3^+^ (Treg) or CD25^-^/FOXP3^-^ (Tconv). (b) Flow cytometric analysis of endometrial tissue single cell preparations from proven fertile women (n=15) showing the proportion of total CD4T cells in the T cell population in the endometrium, and (c) the proportion of Treg in the total CD4T cell population. (d) % CD45RO expression by endometrial (E) (n=12) or peripheral blood (PB) (n=3) Tconv or Treg; bars = median + IQR. (e) Principle Component Analysis (PCA) of RNASeq data from peripheral blood Treg (PB-Tr) and endometrial Treg (E-Treg) or endometrial Tconv (E-Tconv) from nulliparous (Nul) or parous controls (Par), primary RPL (PRPL) or secondary (SRPL). (f) Volcano plot depicting differential gene expression (DESeq2) between endometrial Tconv and Treg, top 25 differentially expressed genes are labelled, (g) heatmap of top 30 upregulated and downregulated genes, red = high, blue = low expression, arrows show genes chose for further analysis in (h) using flow cytometry showing mean fluorescence intensity of intracellular FOXP3 and HELIOS, and cell surface CTLA4, TIGIT, CCR8, CD39 in Tconv (left/grey bars) and Treg (right/blue bars), dots are individual samples and bars = mean+/-S.E.M.

### Phenotypic analysis of endometrial Treg and Tconv cells

We isolated Treg cells (CD3+, CD4+, CD25+, CD127lo cells) or Tconv (CD3+, CD4+, depleted of Treg) from endometrial tissue biopsy digests, from fertile women (nulliparous (n=7) or parous controls (n=3)). In addition we isolated cells from women with primary (n=8) or secondary (n=7) recurrent pregnancy loss (PRPL or SRPL respectively), and Treg were isolated from 5 peripheral blood samples from the RPL patients. RNA-sequencing was performed and principle component analysis revealed that, as expected, endometrial conv-CD4T cells and endometrial Treg cells have distinct signatures, figure 1e. In addition, peripheral blood Treg are separated from the endometrial Treg cluster (figure 1e) indicating an endometrial specific Treg signature.

Differential gene expression between endometrial Tconv and Treg was investigated, depicted by Volcano plot, figure 1f. The gene for the classical Treg signature protein *IL2RA* (CD25) was the most significantly differentially expressed gene, along with other well described Treg genes such as *FOXP3, TIGIT* and *IKZF2 (HELIOS)*. One of the highest differentially expressed genes in endometrial Tconv compared to Treg counterparts was *EOMES*, encoding a transcription factor involved with CD4T cell differentiation. The most abundantly expressed, significantly differentially expressed genes from endometrial Treg cells and Tconv derived from nulliparous control women are shown in figure 1g. Tconv are enriched in genes encoding cytokines (IFN_□_), chemokines (e.g. CCL5 and CCL4), chemokine receptors (e.g. CCR7), cytotoxic factors (GZMA and GZMH) and the anti-inflammatory protein AnnexinA1. In contrast, Treg have higher expression of the regulatory molecules *CTLA4, TIGIT, TNFSRF9* and the chemokine *CCR8*. Along with the classical master Treg transcription factor *FOXP3 gene*, the endometrial Treg express greater levels of the transcription factor genes *BATF, IKZF2* (IKAROS) and *IKZF4* (HELIOS). We confirmed endometrial Treg express significantly higher protein levels of the transcription factors FOXP3 and HELIOS, regulatory molecules CTLA4, TIGIT and CD39 and also the chemokine CCR8, than CD4-Tconv, figure 1h.

### Comparison of peripheral blood and endometrial Treg cells

The peripheral blood Treg were very similar with only 45 genes found to be differentially expressed between the PRPL (n=3) and SRPL (n=2) groups, (DeSeq2 as detailed in Methods, FDR-corrected p<0.01; data not shown). Next we compared peripheral blood Treg cells (n=5) and endometrial Treg cells (n=25, combining data from all controls and RPL patients), a volcano plot of differentially expressed genes with the 25 most significantly altered genes is shown in figure 2a. We found that endometrial Treg have 870 downregulated and 263 upregulated genes compared to peripheral blood Treg (p<0.01 padj and >3, <3 Log2FoldChange ranked on baseMean values), the top 30 up regulated and down regulated genes are shown in figure 2b. We also performed this analysis on the matched samples of endometrial and peripheral blood Treg taken from SRPL patients (n=3), supplementary figure 1, similar genes were found to be upregulated in the endometrium such as *CCR8, CXCR6* and TNF-Receptor superfamily members *TNFRSF-4, −9* and *-18*. Gene Set Enrichment Analysis (GSEA) was performed comparing peripheral blood Treg cells (n=5) and endometrial Treg cells (n=25), the most altered genes between groups are show in Suppl. Figure 2a and pathways enriched by Tregs present in the endometrium compared to peripheral blood Treg elucidated, supplementary figure 2b-c. The key effected pathway involved the receptor signalling pathway, which highlighted IL1 family signalling, Fc receptor binding, and antigen processing along with functions likely to reflect their tissue location such as responses to hypoxia and changes to metabolic processes. Taking this into account, and the DESeq2 results which indicate that many top differentially expressed genes are known Treg markers (such as *ICOS, CXCR6, TNFSRF4, TNFSRF9, TNFSRF18*) this suggests endometrial Treg have a heightened regulatory profile when compared to peripheral blood counterparts. Therefore we focused further studies on proteins involved in immune regulation, tissue residency and antigen presentation. We used flow cytometry to explore the endometrial phenotype. Whilst there was no difference in FOXP3 nor HELIOS expression levels, endometrial Treg expressed higher levels of key regulatory, residency and antigen presentation molecules: CXCR6, ICOS, CTLA4, TIGIT, PD1, IL18R, CD39 and TIM3, but not LAG, than were expressed on Treg in the peripheral blood (p<0.05), figure 2c.

**Figure 2.**
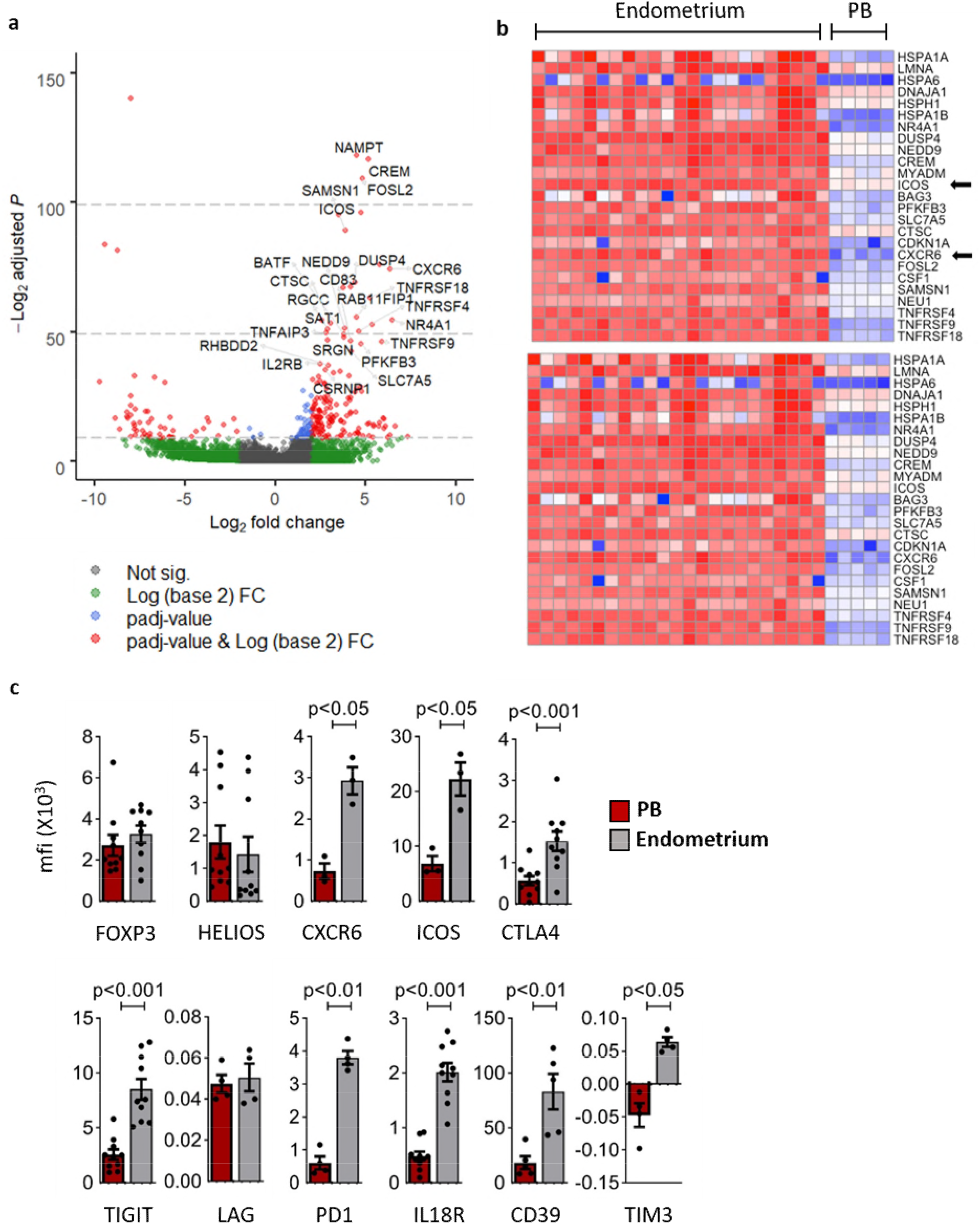
Endometrial and peripheral blood Treg have alternate characteristics. (a) Transcriptomic analysis of Treg derived from endometrium (n=25) versus peripheral blood (PB) (n=5), volcano plot depicting differential gene expression (DESeq2) between endometrial and PB Treg showing top 25 differentially expressed genes, (b) heatmap of top 30 upregulated (top panel) and downregulated (bottom panel) genes in the endometrium versus, red = high, blue = low expression. (c) Flow cytometric analysis of matched peripheral blood (red bars) and endometrium (grey bars) Treg, the mean fluorescence intensity (mfi) of CXCR6, ICOS, CTLA4, TIGIT, LAG, PD1, IL18R, CD39 and TIM3 was determined (bars = mean +/-SEM; dots = individual cell sample).

### Comparison of endometrial Treg and Tconv cells in nulliparous versus parous women

Endometrial Treg or Tconv from controls were segregated into groups of nulliparous (n=6) or parous women (n=3), Treg from parous women had a large number of uniquely expressed genes compared to those from nulliparous women (38.8 versus 1.8%, figure 3a) indicating that exposure to term pregnancy alters the Treg transcriptome. In contrast, the majority of genes expressed in Tconv overlapped (90.7%), with a modest 870 (7.9%) uniquely expressed genes in the parous group, figure 3b. Differential expression analysis yielded 419 upregulated and 801 downregulated genes in Treg from parous women, heatmaps of the top 10 are shown in figure 3c, the highest upregulated gene in the Treg from parous women was IFN□. Comparing nulliparous and parous Tconv differential gene expression using DESeq2 only 17 genes were differentially expressed (data not shown).

**Figure 3.**
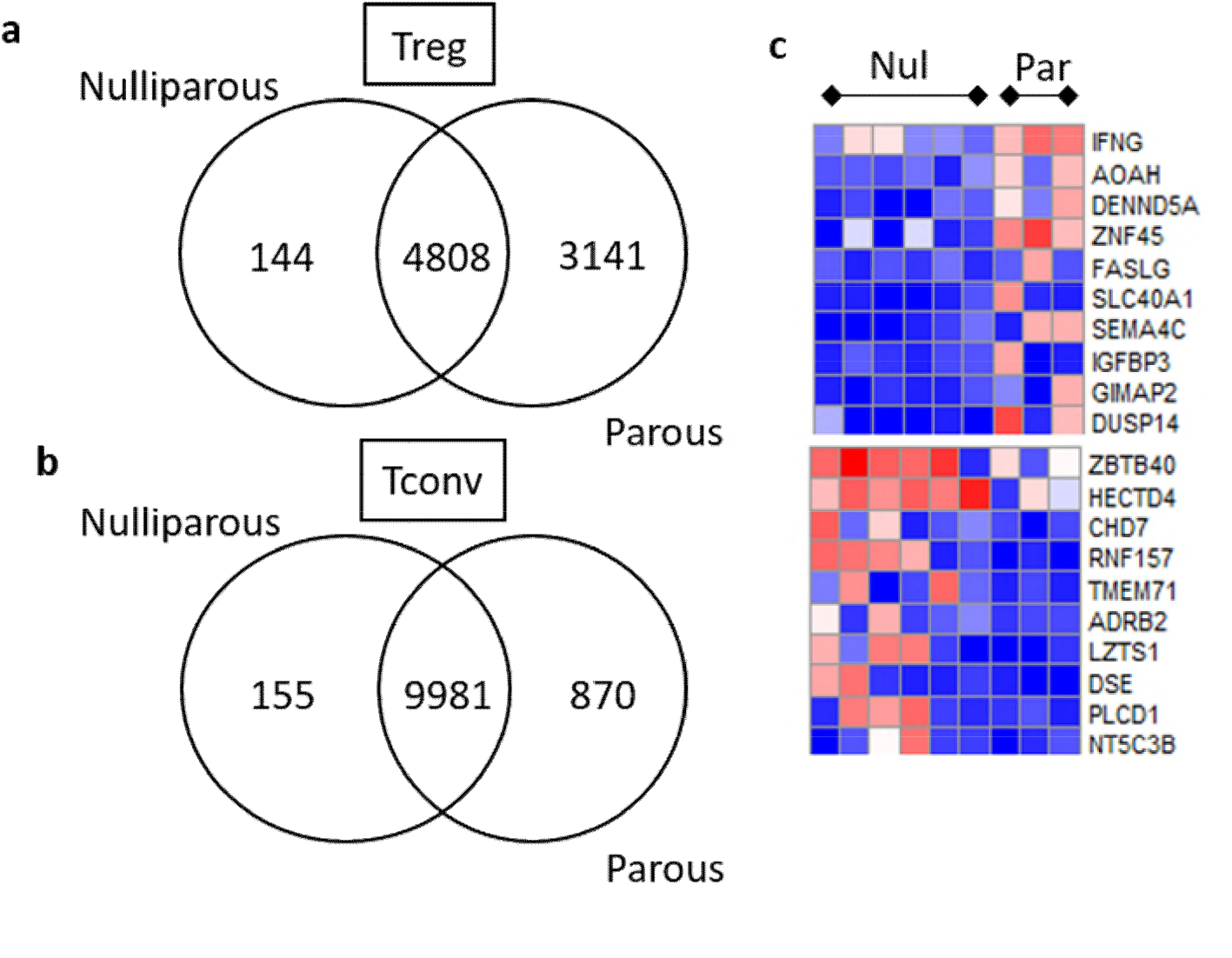
Parous women have an altered Treg, but not Tconv, transcriptome. (a) VENN diagram of uniquely expressed or shared genes within the Treg (a) or Tconv (b) endometrial subpopulations between nulliparous and parous control women. (c) Heatmaps depicting the top 10 most differentially expressed genes between nulliparous and parous women.

### Phenotypic variation of Treg in patients with RPL

As exposure to term pregnancy affected Treg transcription, we next compared cells between nulliparous controls and PRPL and parous controls versus SRPL. First we assessed the proportion of Tconv and Treg within the endometrium between nulliparous and parous controls versus primary and secondary RPL, figure 4a-b. The percentage of CD4Tconv cells in the CD45+ immune cell population was unchanged between groups, similarly PRPL and SRPL did not have altered proportions of Treg within the endometrium. SRPL patients did however have a decreased proportion of Treg when compared to PRPL (p=0.0351), SRPL patients had experienced fewer miscarriages than PRPL (SRPL 3.43+/-0.20) versus PRPL (4.33+/-0.48) (mean +/-1 S.E.M.) however this was not significant and no correlation could be found between the total number of miscarriages experienced and Treg counts, figure 4c.

**Figure 4.**
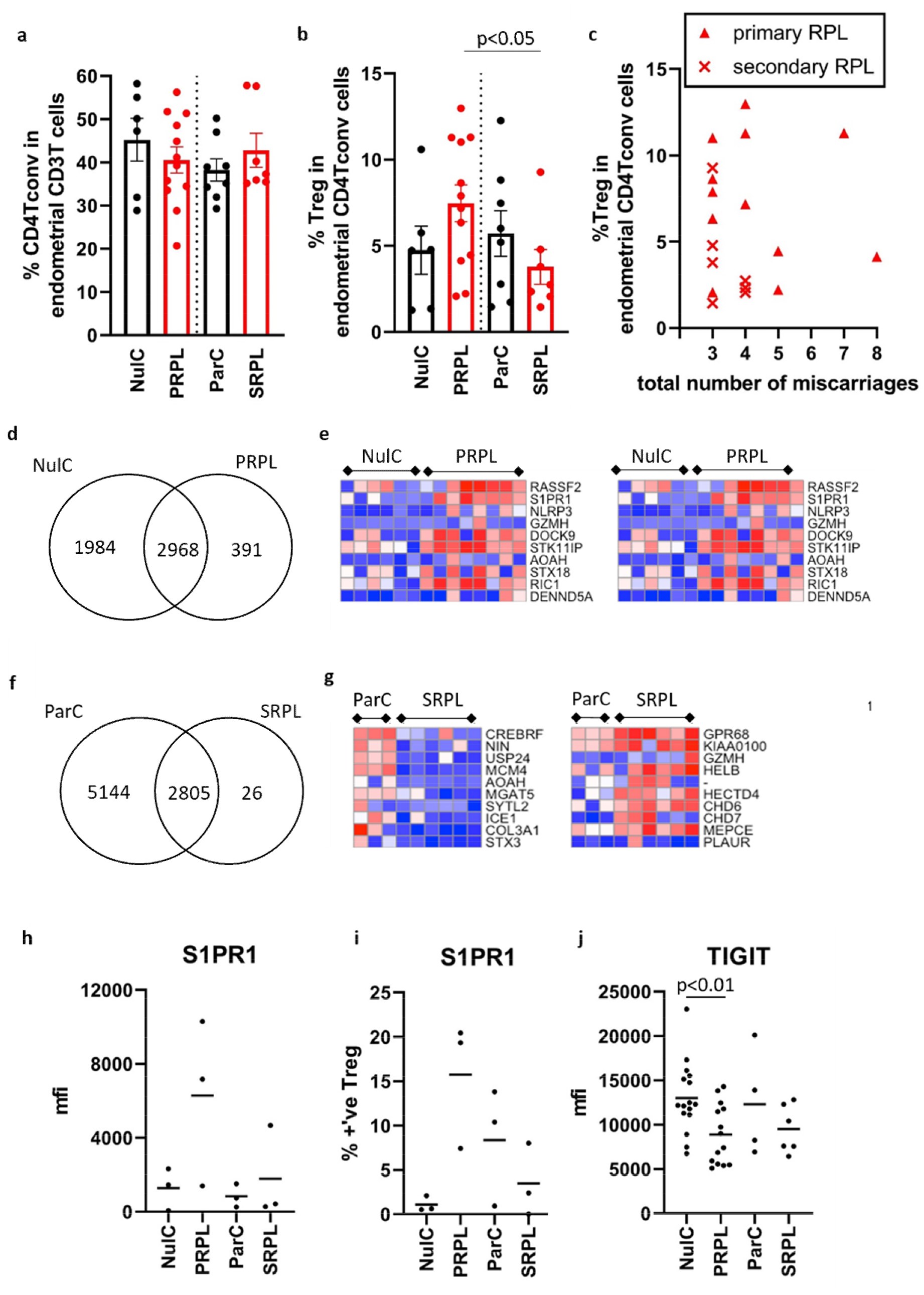
Treg characterisation in Primary and Secondary RPL reveals an altered transcriptome. (a) Tconv and (b) Treg were enumerated by flow cytometry in nulliparous and parous controls (NulC and ParC – black bars) and primary and secondary RPL (PRPL and SRPL – red bars), (c) the Treg proportion of the CD4 Tconv population versus the number of miscarriage in PRPL (triangles) and SRPL (crosses). (d) were compared analysing uniquely expressed and shared genes and top 10 differentially upregulated and downregulated expressed genes are displayed as a heatmap between (e) NulC and PRPL Treg or (g) ParC and SRPL. Flow cytometric analysis of cell surface (h-i) S1PR1 and (j) TIGIT between RPL and control patients showing mean fluorescent intensity (mfi)(h and j) or the % positive Treg cells (i) detected by flow cytometry. Bars = mean value.

Next we compared gene signatures of endometrial Treg, comparing PRPL to nulliparous controls, the majority of genes overlapped, however unique genes were identified in nulliparous controls and PRPL, figure 4d. Differential gene expression analysis identified 657 upregulated and 716 downregualted genes, the top 10 up- and down-regulated genes are displayed as heatmaps in figure 4e.

Comparing SRPL to parous controls, few genes were unique to SRPL compared to the parous controls, figure 4f, however 1466 genes were lower and 773 genes higher in expression in SRPL when compared to parous controls, top 10 differentially expressed genes are shown in figure 4g. Of note, all comparisons were also performed on Tconv, PRPL and SRPL had similar transcriptomic profiles to their controls, with 45 and 79 total differentially expressed genes between groups respectively (data not shown).

The second highest upregulated gene in PRPL was *S1PR1*; Sphingosine-1-Phosphate receptor-1 is a G-Protein coupled receptor involved with lymphocyte activation, migration and trafficking (13). We analysed mRNA expression of this factor, along with other factors identified by DESeq2 between groups and also known Treg factors identified as core signature genes, such as transcription factors, regulatory molecules, tissue residency/chemoattraction/exit molecules (14), between all Treg and Tconv cell subsets, supplementary figure 3. Endometrial Tconv have higher *Tbet* and *RORyc* than Treg, whereas endometrial Treg have greater *FOXP3, HELIOS* and *BLIMP1. CTLA4, TIGIT* and *CD39* were greatly enhanced in endometrial Treg compared to Tconv and also peripheral blood Treg. *S1PR1* was higher in PRPL Treg than both control groups and also SRPL (2/6 expressed *S1PR1*) but genes for other tissue residency factors such as *CD103* or the linked *CD69* did not follow this trend. *CCR7* and *CXCR3* are likely Tconv chemoattraction markers, whereas *CXCR6* and *CCR8* were higher in Treg. We further analysed protein expression levels of several of these identified factors between controls and RPL. PRPL patients had higher levels of Treg S1PR1 than both control groups and SRPL patients, confirmed by flow cytometry, when considering both the mean fluorescence intensity of expression and the proportion of Treg cells present expressing S1PR1, however due to small sample sizes statistical analysis is not performed, figure 4h and 4i. In addition, cell surface TIGIT had significantly lower expression on Treg derived from PRPL patients than nulliparous controls, figure 4j, even though this factor was not differentially regulated at the transcriptomic levels. Of note FOXP3, HELIOS and CTLA4 protein levels were unchanged between groups (data not shown).

In conclusion, we have identified that endometrial and peripheral blood Tregs have altered regulatory transcriptomic and protein signature profiles, and that these profiles are altered by exposure to term pregnancy. In addition, we have identified not only changes to Treg between RPL and controls at the transcriptomic level, but also provide evidence of post-translational proteomic changes to Treg in RPL patients that would lead to potential Treg change of function.

## Discussion

The endometrial CD4T cell compartment occupies a lower proportion of the total CD3+ cell population than that in the PB; however 5% of CD4+T cells are FOXP3+/CD25hi/CD127lo Treg both in the endometrium and PB, a similar Treg proportion was reported in other mucosal tissues isolated from adults (15). RNA sequencing analysis of endometrial CD4+ Tconv and Treg cells was performed and differential gene expression analysis highlights the core transcriptome, figure 1. Tconv express the genes for granzyme A and H and IFN□ suggesting the presence of a population of cells with pro-inflammatory/cytolytic responses, possibly driven by the highly differentially expressed transcription factor EOMES in Tconv which is synonymous with this phenotype (16). Tconv also expressed higher levels of *CCL4* (MIP1β) and *CCL5* (RANTES) indicative of capacity to recruit immune cells to the endometrium, which help facilitate embryo implantation through tissue remodelling (17). Endometrial Treg, sorted for sequencing using CD25hi/CD127lo expression, displayed *CD25* (*IL2RA*) as the highest transcriptionally differentially expressed gene between Treg and Tconv, other highly enriched Treg specific genes were also expected, such as the transcription factor *FOXP3* and the regulatory molecules *CTLA4, TIGIT and CD39*, which were also validated via flow cytometry. CCR8 expression was highly enriched in Treg, the CCL1-CCR8 axis is known to potentiate Treg suppressive function (18) which is in line with the phenotype noted here.

We targeted key T cell transcription factors (supplementary figure 3) and found that Treg were also high for *GATA, BLIMP1, STAT3*. The Tconv population contained cells expressing *Tbet, GATA* and also some *RORC*, the Tconv population are therefore as expected a likely combination of Th1, Th2 and Th17 cells. In addition, Treg expressed *IKZF2* (*HELIOS*) was also detected; HELIOS, along with Neuropilin, has been suggested by some researchers to be expressed by peripheral Treg (pTreg) as opposed to thymic derived Treg, however controversy exists as to their true value as such markers (19), here we find low endometrial Treg gene transcripts for Neuropilin and higher HELIOS with variable protein levels.

Principal component analysis, figure 1e, separated PB and Endometrial Treg into distinct clusters, therefore we compared the endometrial Treg (all subgroups) to PB Treg, figure 2. This analysis was also performed with the matched endometrial and PB Treg from 3 specific SRPL patients, supplementary figure 1. An endometrial specific tissue Treg signature has not yet been described, but it is to be expected as Tregs have non-lymphoid tissue specific local adaptations (14). CXCR6 was highly expressed by endometrial Treg, this has previously been shown to be upregulated in human colon Treg (14). ICOS (inducible T-cell costimulator) was also highly upregulated in endometrial Treg, *in vitro* ICOS+ Treg produce abundant IL-10 (20) and ICOS expression is closely linked to Blimp1 expression in Treg located in mucosal sites (21) as we have detected in these endometrial Treg. Immune-checkpoint molecules are a key target for immunotherapy, upon ligation inhibitory signals limit immunity (22); family members include cytotoxic T-lymphocyte-associated protein 4 (CTLA-4), T-cell immunoreceptor with immunoglobulin and ITIM domains (TIGIT), programmed cell death protein 1 (PD-1), T-cell immunoglobulin and mucin-domain containing-3 (Tim-3) and lymphocyte-activation gene 3 (LAG-3), all of which have been studied here. Of note the PD-1/PD-L1 pathway is critically important in murine pregnancy as blockade causes fetal resorption, due to an imbalance in Th17/Treg cells which is diminished with Treg cell adoptive transfer (23). The phenotype identified here indicated endometrial Treg have enhanced expression levels of several immune checkpoint molecules, indicating endometrial Treg cells have differing phenotypic functions to those found in the blood.

This is the first description of the CD4T and Treg compartment in human endometrium comparing parity and controls. We know that Treg have fundamental importance for materno-fetal tolerance: CNS1 dependent extrathymic differentiation of Treg occurred concurrently with placental evolution (24), and in mice Tregs expand during pregnancy forming a memory response that is recalled in subsequent pregnancy (25). Previous studies on decidua tissue taken from term placenta samples show that Tregs undergo clonal expansion throughout pregnancy, of note this cannot be detected in the peripheral blood Treg (26), but peripheral blood Treg numbers increase in human pregnancies as gestation progresses. Our observation that the CD4Tconv compartment is similar between the PB and endometrium, whereas Treg have greater significant phenotypic differences, suggests Treg tissue specificity, this was observed by a study analysing the T cell receptor beta variable (TRBV) repertoire that found most variation in the Treg population between cells derived from the blood or decidua (27). IFN□ was identified as the most upregulated gene in Treg from parous women. Interestingly, parous women have been shown to harbour a subpopulation of NKG2C^hi^ dNK cells thought to be produced during their first pregnancy, which also have increased capacity to produce IFN□ along with VEGF□, both are known regulators angiogenesis and the vascularisation required for efficient placentation (28). Future work to assess if Treg from decidua secrete IFN□ and contribute to placentation is required. Indeed Treg are altered during pregnancy: decidual Treg have the ability to supress both fetus-specific and non-specific responses (29, 30) and Treg (FOXP3+CD25hi) in first trimester decidua, compared to peripheral blood counterparts, express higher CCR5, ST2, CD25, BATF, IL10, GITR, GARP and CCR8 (31). Here we describe that prior to pregnancy, the endometrial Treg already express this enhanced regulatory phenotype, and that changes to the population in pregnancy are long lived in the endometrial tissue.

Given therefore that the exposure to pregnancy has a significant impact on endometrial Tregs, we compared nulliparous women to patients with PRPL, and correspondingly parous women to SRPL. Using flow cytometry to quantify the proportions of Tconv in the T cell population and Treg in the Tconv populations, we found no significant differences in the cell proportions between groups (fig. 4b). Treg/CD4 Tconv cell proportions in human endometrium have not been previously clearly defined (32), there is limited evidence of increased FOXP3+ cells (immunohistochemical analysis) in the endometrium of RPL patients (33), but Tconv and Treg proportions and populations have mainly been studied in in the peripheral blood or decidua obtained after miscarriage. Fertile women have expanded peripheral blood CD4+CD25+FOXP3+ Treg in the late follicular phase, this expansion is not seen in RPL which also have reduced functional capacity to inhibit proliferation (34, 35) and inhibit NK cell cytotoxicity (36). In general, CD4T cells numbers are lower in RPL decidua than controls (37) and in normal early pregnancy Tconv were not clonally expanded (38). CD4^+^CD25^bright^ cells were a higher proportion of the CD4T cell compartment in early pregnancy decidua obtained after induced abortion (21.84 ± 2.92) than after sporadic miscarriage (7.14 ± 1.85) (39), and Treg numbers are reportedly reduced in decidua from RPL patients compared to normal first trimester pregnancy (35, 40-42). Here we find that the numbers of cells present are similar prior to pregnancy, in RPL and controls, therefore the observed reduction in Treg numbers in decidua in women suffering miscarriage is likely a consequence of failure to recruit or expand Treg populations upon establishment of pregnancy. Nevertheless we do observe phenotypic differences in the cells, prior to pregnancy, as discussed below.

Treg derived from PRPL had altered Rassf2, a novel tumour suppressor gene with associated K-Ras pro-apoptotic effector functions and the second highest differentially expressed gene was S1PR1, figure 4. S1PR1 has well established roles in T cell trafficking out of lymphoid tissue, it’s role for T cell egress from mucosal tissues is less well defined (13) and little is known about S1PR1 expression on human Treg. Mice with S1PR1 deficient Treg develop autoimmunity, with Treg displaying an activated phenotype prone to apoptosis (43). We chose to study Treg S1PR1 cell surface expression further; flow data supports the RNAseq analysis revealing PRPL patients had higher expression levels than controls, figure 4. The functional role of endometrial S1PR1 Treg expression remains to be determined, and if the expression indicates a subset of Treg in PRPL which can exit the tissue. In addition, we also studied key Treg immune checkpoint molecules, and found that endometrial Treg from PRPL had significantly lower TIGIT protein expression levels that corresponding controls. This is the first observation of altered Treg populations in RPL, phenotypic changes indicate reduced inhibitory capacity in PRPL. Whether patients with PRPL would benefit from therapies enhancing tolerance through this pathway merits further investigation.

## Methods

### Tissue Collection and Processing

The study was approved by the Oxford Research Ethics Committee C (ref:08/H0606/94). All participants gave written informed consent in accordance with the Helsinki Declaration of 1975.

All women were <40 years of age. Blood or endometrial samples were taken in the mid-luteal phase of the menstrual cycle. Participants were recruited at least three months post miscarriage or hormonal treatment. Women included in this study had experienced at least 3 consecutive miscarriages. All women with RPL had normal thyroid function, negative anti-phospholipid screen (cardiolipin IgG Ab, anti-beta-2 glycoprotein 1 IgG and DRVVT ratio), negative thrombophilia screen (including Factor V Leiden, Prothrombin 20210 mutation, Antithrombin III, protein C, protein S) and no evidence of uterine structural abnormalities (identified by ultrasound scan or hysteroscopy). Maternal and paternal karyotypes were only carried out if an unbalanced translocation was identified in karyotyped miscarriage tissue as per the Royal College of Obstetrics and Gynaecologists guidelines (44). Control endometrial samples were taken in the cycle prior to IVF. Controls for SRPL patients were taken from women who had previous live birth pregnancy outcomes. Women had experienced none or no more than 1 miscarriage and patients with endometriosis or known autoimmunity were excluded. Control samples for RNAseq or matched control to RPL flow cytometry experiments were from fertile patients who either have had a previous live birth but were undergoing IVF for male factor infertility (Parous Controls; ParC), or a who had a live birth after the IVF cycle following endometrial biopsy (Nulliparous Controls; NulC).

Endometrial samples were obtained using an Endocell disposable endometrial cell sampler (Wallach Surgical devices, CT, USA) and digested using 1X Liberase (Roche Life Sciences) as previously described (7). Peripheral blood was collected into sodium heparin anti-coagulant (10 U/ml) and peripheral blood mononuclear cells (PBMC) isolated using lymphoprep (Axis Shield Diagnostics). Single cell suspensions were frozen in 10% DMSO/ FCS using a Nalgene Mr. Frosty Freezing chamber (Thermo Fisher), before being transferred to liquid nitrogen for storage prior to use.

### RNA Sequencing patient sample information

Matched blood and endometrial samples (n=5) were taken from women with an average age of 36.2 +/-4.1 years (Mean, S.D.), 2 were nulliparous, 4 patients had experienced 3 miscarriages and one patient had 7 prior miscarriages. Details of patients that provided endometrial samples for RNAseq are shown in Table 1.

**Table 1:**
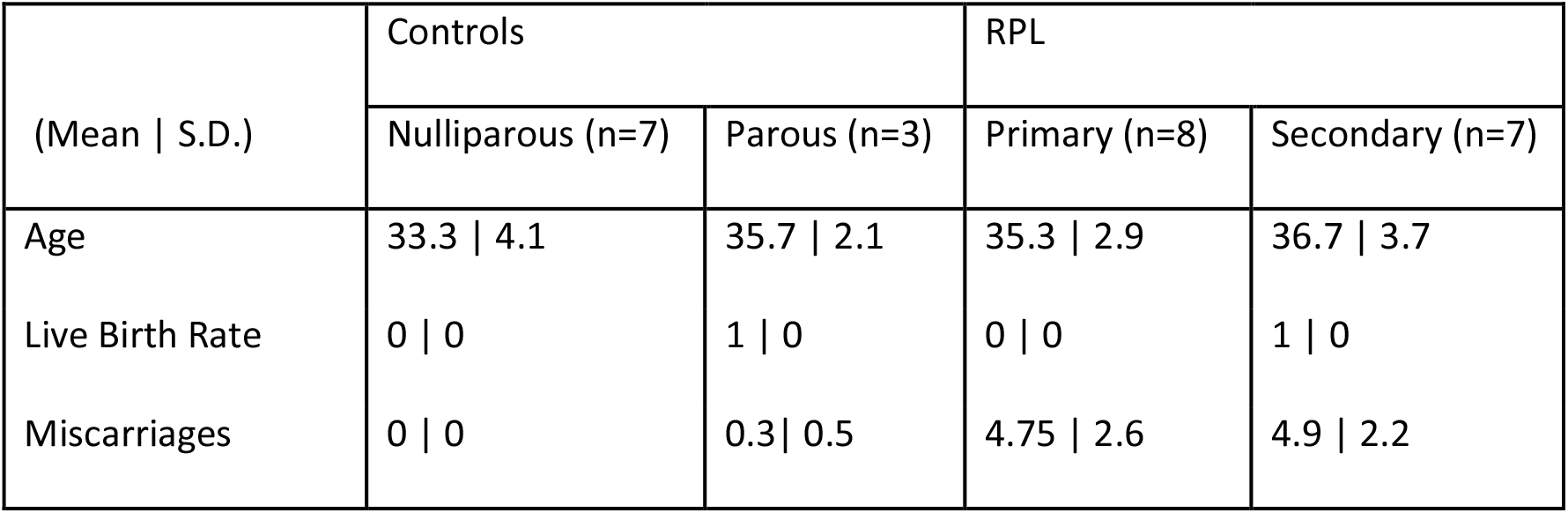
Details of patients that provided endometrial samples for RNAseq.

| (Mean \| S.D.) | Controls |  | RPL |  |
| --- | --- | --- | --- | --- |
|  | Nulliparous (n=7) | Parous (n=3) | Primary (n=8) | Secondary (n=7) |
| Age | 33.3 \| 4.1 | 35.7 \| 2.1 | 35.3 \| 2.9 | 36.7 \| 3.7 |
| Live Birth Rate | 0 \| 0 | 1 \| 0 | 0 \| 0 | 1 \| 0 |
| Miscarriages | 0 \| 0 | 0.3 \| 0.5 | 4.75 \| 2.6 | 4.9 \| 2.2 |

### Cell Sorting for RNAseq and RNA extraction

Frozen endometrial cell digests or isolated PBMC were thawed in to HS-media (RPMI1640 supplemented with 10%FCS, 1% Human Serum, glutamine and pen/strep) at 37□C and centrifuged at 300 Х g. Endometrial cells were resuspended in HS-Media and transferred to a 25cm^2^ flask for 30 minutes at 37□C/ 5% CO_2_, then supernatants containing non-adherent cells were further transferred to a fresh flask for an additional 30 minutes incubation, for stromal and epithelial cell depletion. Supernatants were then removed and transferred onto a lymphoprep layer with centrifugation at 800 x g for 20 minutes. Cells in the interface were washed in PBS and prepared for cell sorting on an Aria III (BD Biosciences). Briefly, both PBMC or endometrial cells were stained with Zombie Aqua™ Live/Dead Viability dye then incubated with antibodies (CD3-APC Fire750, CD45-Briliant Violet (BV)650, CD4-FITC, CD25-PE, CD8-BV711, CD19-PeCy7, CD127-APC), all reagents are from Biolegend unless otherwise stated.

CD4T cells were isolated as singlet cells (FSC-H vs FSC-A), Zombie Aqua-, CD45+, CD3+, CD4+, CD8-, CD19-cells and Treg were Zombie Aqua-, CD45+, CD3+, CD4+, CD8-, CD19-, CD25+ and CD127lo. Immediately after sorting, cells were pelleted by centrifugation and resuspended in RLT (Qiagen) and RNA was extracted using QIAGEN RNeasy Micro Kits according to the manufacturer’s protocol. RNA quantity and quality were accessed on Agilent 4200 TapeStation System.

Sorted cell populations (mean +/-1.S.D.) were 2555 +/-2254 for endometrial Treg, 36373 +/-37819 for endometrial CD4T cells and 34450 +/-14531 blood Treg. No significant differences were detected within sorted cell population counts between nulliparous controls, parous controls, primary RPL and secondary RPL.

### Library preparation and RNA sequencing

Samples were processed following Smart-Seq 2 protocols (45), the library was prepared using a Standard Nextera Illumina Library Prep kit using a unique dual-index strategy to barcode cDNA fragments. DNA samples were amplified for 16 cycles and the cDNA quantities assessed using a 2100 Bioanalyzer (Agilent, USA) with Quant-iT PicoGreen (Thermo Fisher, USA). Clustering and sequencing was carried out by Novogene Co., Ltd. Briefly, clustering of the index-coded samples was performed on a cBot Cluster Generation System using PE Cluster Kit cBot-HS (Illumina) and paired end RNA sequencing was performed using the Novoseq 6000 platform, with 125 bp/150 bp read length and approximately 25 million reads per sample. Quality control was performed by Novogene Co., Ltd and reads mapped to Ensemble Homo Sapiens cDNA database release 94. 1 NulC and 1 SRPL Treg sample did not pass QC and were excluded from analysis.

### Data Analysis

Data analysis was performed using R (version 3.5.3) and RStudio, differential gene expression analysis was performed using DESeq2 (46). Heatmaps were generated using p<0.01 padj and >3, <3 Log2FoldChange values and top 30 changes in expression value shown ranked by baseMean expression levels. PCA plots were generated using vst transformation. Gene Set Enrichment Analysis (GSEA) (47) was performed using GSEA software (version 4.0.2) and results visualized in Cytoscape software (version 3.7.2) (48). Venn Diagrams were generated using genes with reads >10 and calculated using Bioinformatics & Evolutionary Genomics tool hosted by the University of Ghent, Belgium (http://bioinformatics.psb.ugent.be/webtools/Venn/)

### Flow Cytometry Phenotyping

All flow cytometry reagents were from Biolegend unless otherwise stated. Samples were incubated with Zombie-Aqua Fixable Viability kit for 15 minutes in the dark, then cells washed in PBS/2%FCS and antibodies towards cell surface markers were added for 20 minutes at 4□C in the dark. Antibodies used were CD4-FITC(OKT4), CD8a-PE(HIT8a), CD8-PeCy7 (RPA-T8), CD3-PeCy5(UCHT1), CD56-PeCy7(BD Biosciences; B159), CD16-APCCy7(BD Biosciences; 3G8), CD45-AlexaFluor700 or CD45-APC (HI30),CD45RO-APC-Cy7(UCHL1), CD127-AlexaFluor647(A019D5), PD-1(CD279)-APC(EH12.2H7), CXCR6-PECy7 (K041E5), CD25-PE/Dazzle594 (M-A251), CCR8(CD198)-PE (L263G8), CD3-APC/Fire750 (SK7), CD45-AlexaFluor700 (HI30), CD127-BV711 (A019D5), ICOS-BV650 (C392.4A), CTLA-4(CD152)-BV786 (BD Biosciences; BNI3), TIGIT-BV421, (A15153G), ENTPD1(CD39)-BV421 (xxx), LAG-3(CD223)-PE (113C65), TIM-3(CD366)-BV785 (F38-2E2), IL18R-PE (H44), S1PR1-eFluor550 (eBiosciences; SW4GYPP). Afterwards, cells were washed in PBS/2% FCS and intranuclear antibody staining was performed using ‘True-Nuclear Transcription Factor Buffer Set’ (Biolegend), cells were fixed for 1 hour then transferred into PBS/ 2% FCS overnight, then permabilisation performed before addition of antibodies FOXP3-AlexaFluor647 (259D), IKZF2(HELIOS)-PeCy7 (22F6) and additional CTLA-4 antibody for total cellular CTLA-4 measurement, for 45 minutes incubation.

Data was acquired using a LSR-II flow cytometer (BD Biosciences) and data analysed using FlowJo software (Tree Star Inc.), fluorescence Minus One (FMO) controls established gating strategies. Graphs were plotted and statistics generated in GraphPad Prism 8.4.3.

## Data Availability

Data is available upon request

## Acknowledgments

The authors would like to thank the participants in this study, and the staff at Oxford Fertility, the John Radcliffe Hospital and University Hospitals Coventry and Warwickshire and Warwick Medical School who have assisted with the collections of samples. Dr Helen Ferry, NDM University of Oxford assisted with flow cytometric sorting and Dr Neil Ashley, WIMM University of Oxford assisted with the SmartSeq2 and library preparation. Funding was from the IVI Foundation FINOX.

## Author Contributions

JS and IG conceived of the study. Data was generated and analysed by JS, MS, HRC, GC, GM, EO’D and ED. JB, SQ, TC and IG provided clinical expertise to the study design. All authors contributed to refinement of the study protocol and approved the final manuscript.

---

**Supplementary Figure 1.**
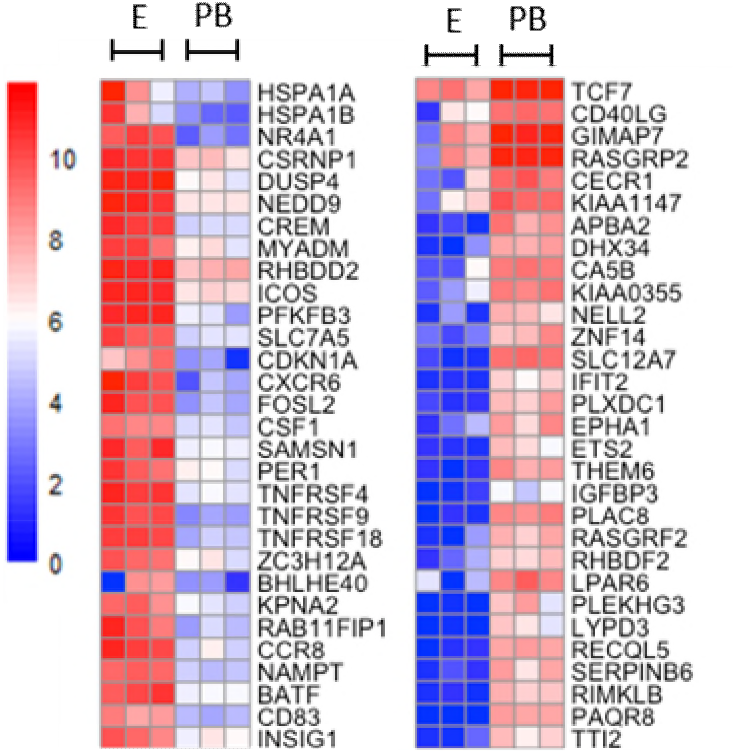
Transcriptomic analysis of Treg derived from matched endometrium (E) (n=3) versus peripheral blood (PB) from SRPL patients (n=3), in total 571 genes were upreguiated and 169 genes were downregulated. Heatmap of top 30 upregulated (top panel) and dowmegulated (bottom panel) genes in the endometrium versus PB, red = high, blue = fow expression.

**Supplementary Figure 2.**
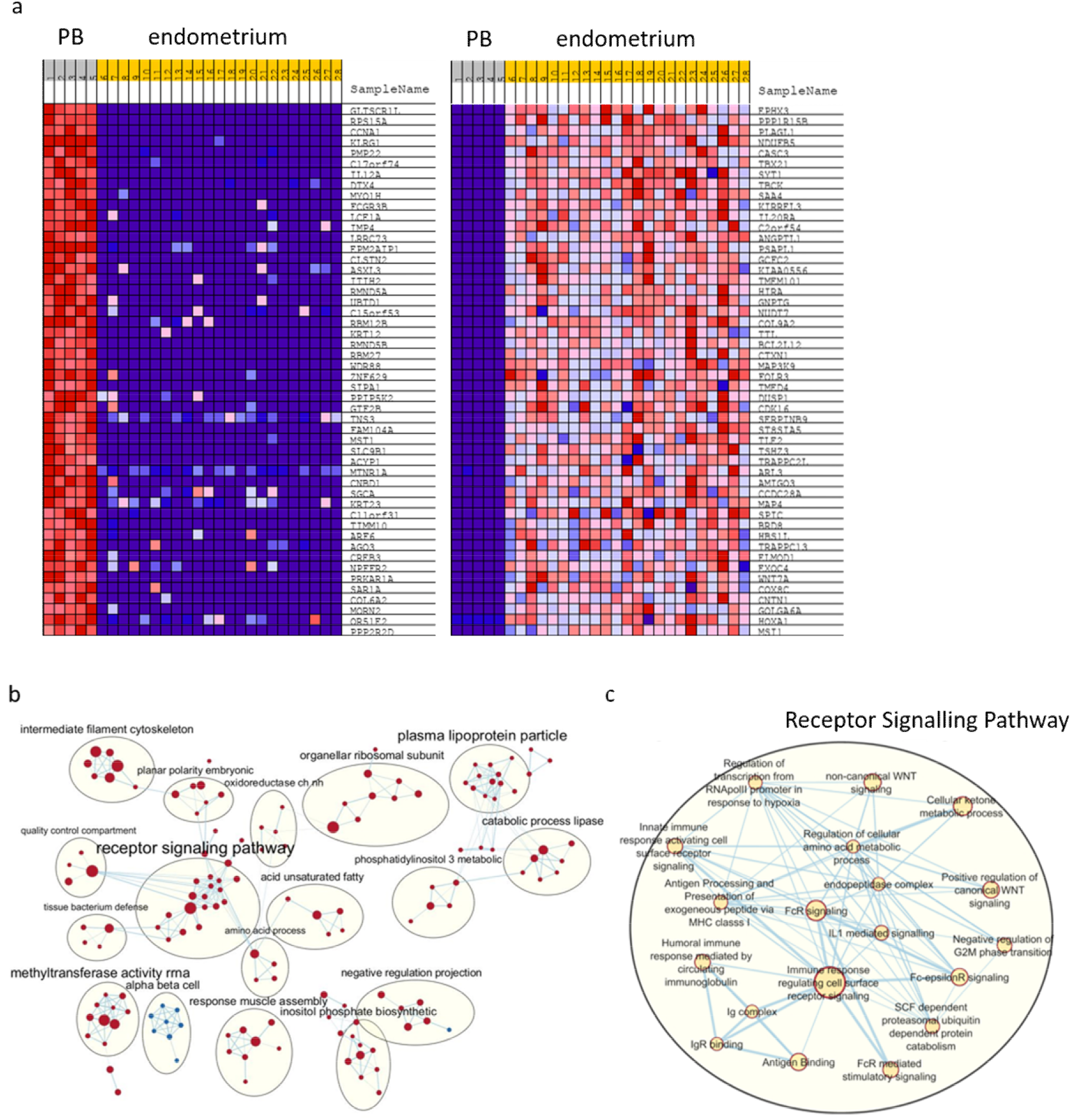
Gene set Enrichment Analysis (GSEA) (Broad Institute), was performed using the RNA transcriptome from peripheral blood (1-5) or endometrial (6-28) Treg. The Gene Ontology gene set database C5.all.v7.0 was used with 1000 permutations, leading edge analysis performed and Enrichment Map calculated with P-value cutoff 0.0005 and FDR Q-value cutoff 0.1, figures were visualized using Cytoscape 3.7.2 with ≥ nodes selected. a) heatmap of top 50 altered genes between groups (red – high expression, blue – low expression), b) pathways enrichment map, and c) enlargement of the highest implicated pathway’ receptor signalling pathway’.

**Supplementary Figure 3.**
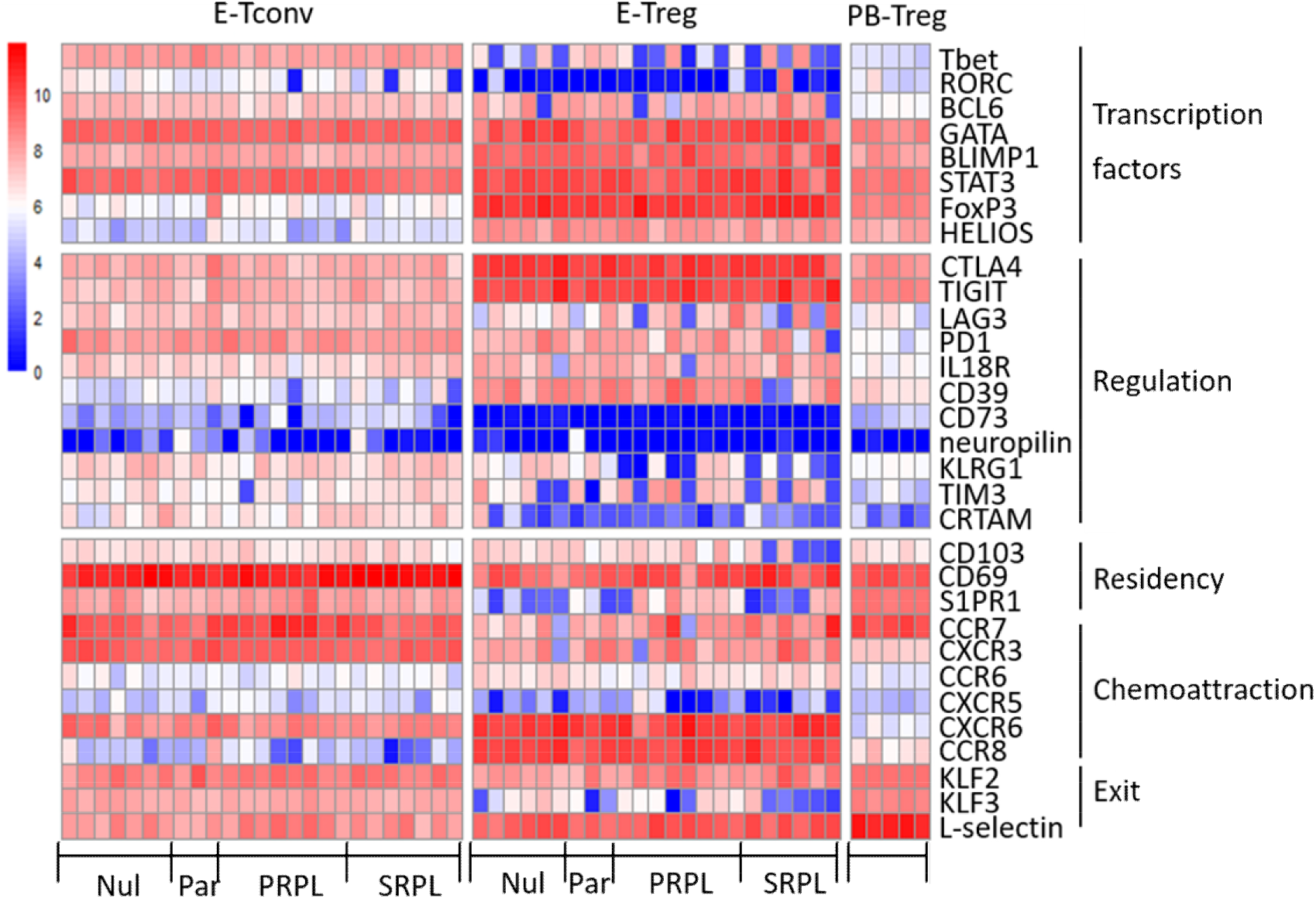
Transcriptomic analysis of Treg derived from matched endometrium Tconv (E-Tconv) and Treg (E-Treg) (n=25) versus peripheral blood Treg (PB-Treg) (n=5) from Nulluparous (Nul), Parous (Par), Primary RPL (PRPL) and Secondary RPL (SRPL) patients, heatmap of gene expression levels of known Tconv and Treg factors, red = high, blue = low expression

